# Understudied anophelines may sustain residual transmission during the dry season in a pre-elimination setting in southern Zambia

**DOI:** 10.1101/2025.03.31.25324992

**Authors:** Anne C. Martin, Victoria Kamilar, Limonty Simubali, Twig Mudenda, Harry Hamapumbu, Jessica L Schue, Mary E. Gebhardt, Reneé L.M.N. Ali, Jennifer C. Stevenson, Timothy Shields, Michael R. Desjardins, Frank C. Curriero, William J. Moss, Douglas E. Norris

**Affiliations:** Department of Epidemiology, Johns Hopkins Bloomberg School of Public Health, Baltimore USA; The Johns Hopkins Malaria Research Institute, Johns Hopkins Bloomberg School of Public Health, Baltimore, MD, USA; Macha Research Trust, Choma District, Zambia; Department of International Health, Johns Hopkins Bloomberg School of Public Health, Baltimore USA; The W. Harry Feinstone Department of Molecular Microbiology and Immunology, Johns Hopkins Bloomberg School of Public Health, Baltimore USA; Spatial Science for Public Health Center, Johns Hopkins Bloomberg School of Public Health, Baltimore, Maryland, USA; Department of Oncology, Johns Hopkins School of Medicine, Baltimore, Maryland, USA

**Author notes:** **Corresponding author contact information:** Anne Martin, 615 N Wolfe St, Baltimore MD, 21205, USA.

## Abstract

Malaria control is a public health priority but common control methods like indoor residual spraying and the use of bednets do not target outdoor-biting vectors. In settings with seasonal residual malaria transmission, we lack critical knowledge regarding anopheline species composition and their role in transmission. This study aimed to determine relative seasonal vector species abundance and associated household level factors in a low transmission setting in Choma District, Zambia. Indoor and outdoor adult vector collections were embedded in a community-based longitudinal cohort study in 60 households that were visited monthly for 2 years between 2018 and 2020. Surveys conducted at the time of trap placement collected information on animal ownership, housing structure, and the receipt of malaria interventions. Anopheline species identities were molecularly confirmed by polymerase chain reaction, and enzyme-linked immunosorbent assay was used to detect the circumsporozoite protein of *Plasmodium falciparum*. Generalized linear mixed effects negative binomial regression with zero-inflation models were used to describe the relationship between risk factors and the outcome of monthly anopheline counts at each household, stratified by season. The study collected 1,532 female anophelines, 76% of which were caught outdoors. The relative abundance differed by season: in the dry season, 90% of female anophelines were caught outdoors. *Anopheles arabiensis* was overall the most common vector, but made up only 28% of outdoor collections; the remainder were understudied anophelines including *An. coustani*, *An. leesoni*, *An. rufipes*, *and An. squamosus.* The only *Plasmodium falciparum*-infected mosquito was an *An. squamosus* that was caught outdoors. Owning more goats was associated with a 3.5 (IRR 4.47, 95% confidence interval [CI]: 2.00, 10.01) and 7.7 (IRR 8.73, 95% CI: 4.40, 17.32) times increase in indoor and outdoor anopheline collections in the dry season and a 1.2 (IRR 2.18, 95% CI: 1.12, 4.23) times higher risk of outdoor anophelines in the rainy season. Improved housing structure was associated with fewer indoor anophelines in the rainy season, but not during dry season or outdoor anopheline abundance any time of year. Vector control in this low transmission setting, therefore, needs to target anopheline mosquitoes year-round, must be expanded to target traditionally zoophillic mosquitoes, and leverage known risk factors when selecting methods of control.

## Introduction

With a global burden of 263 million cases in 2023, malaria and its control remain a public health priority (1). While 26 countries have successfully eliminated malaria this century, countries in sub-Saharan Africa with highly heterogenous malaria transmission, which include low transmission “pre-elimination” regions, may require different approaches to achieve and maintain elimination locally (2). The classic vector control methods used to curb malaria transmission in high transmission settings are indoor residual spray (IRS) and the use of long-lasting insecticide treated bednets (LLINs). Malaria transmission requires human-vector interaction, and the effectiveness of these vector control methods depends on whether this interaction is interrupted.

Vector behaviors such as host preference, foraging location and time, and resting area preference influence the likelihood and duration of these interactions and the effectiveness of existing vector control. *Anopheles gambiae* sensu stricto (s.s.), *An. coluzzii* and *An. funestus* s.s., principle vectors of African malaria, are highly anthropophilic and prefer to feed and rest indoors, and thus indoor vector control methods like IRS and bednets work well to protect humans (3). In contrast, other well-studied malaria vectors like *An. arabiensis* demonstrate variability in their anthropophily (4). The plasticity of their behaviors, at times being exophagic and exophilic, may allow them to evade the killing and repellency effects of IRS and bednets (5,6). Other species, such as *An. squamosus*, *An. rufipes*, and *An. coustani* demonstrate less stringent host preferences and exhibit more zoophilic, exophagic, and exophilic biting and resting behaviors (7–9). However, there are documented examples, such as in Madagascar, where these long-overlooked anopheline species can become major malaria vectors, with their foraging and resting behaviors largely unimpacted by indoor-based vector control methods (10).

The selection of vector control tools must consider species composition and, when possible, their distribution should target households with risk factors for higher vector abundance. While there is ample literature on risk factors for malaria infection, there are fewer studies explicitly focused on risk factors for household vector burden in low transmission settings. In high transmission settings, indoor vector control, house construction materials, screenless windows, open eaves, household elevation and distance from water bodies, rainfall, vegetation index, and presence of livestock are all correlated with indoor vector density (11–13). In low transmission settings, the importance of these factors may differ. For example, microclimate may drive local epidemiology in low transmission settings as conditions surrounding a given household may have greater influence on vector abundance than in high transmission settings where there are ample vector breeding sites distributed throughout the landscape (14). Additionally, settings of low and residual transmission may be the result of changes in local ecology and climate that support a shift from common vector species (e.g. *An. gambiae, An. arabiensis*) to more cryptic species such as *An. squamosus* with different or unknown bionomics and behaviors (15–17). Indoor-based vector control measures may be less effective if species are more exophagic and exophilic, and animal ownership may be more important if species tend to be zoophilic.

This study aimed to determine relative vector species abundance and identify household level factors associated with anopheline abundance in a low transmission setting in Choma District, Zambia. We conducted indoor and outdoor adult vector collections that were embedded in a two-year community-based longitudinal cohort study. We hypothesized that there would be no protective association between mosquito abundance and IRS or use of bednets, but that there would be an association with other household and environmental factors.

## Material and Methods

### Ethics statement

This study was a part of the “Against Transmission Of Malaria With Everyone (ANTOOMWE) study,” which had ethical approval from the Johns Hopkins Bloomberg School of Public Health (Baltimore, Maryland) under IRB no: 00003467 and the Tropical Diseases Research Center (TDRC under IRB no: TDRC/ERC/2010/14/11. Written consent was obtained from an adult household representative to enroll the household in monthly surveillance. Oral consent was obtained for each month of entomological surveillance.

### Study site and data collection

The cohort study took place from 2018 – 2020 in the catchment area of Mapanza Rural Health Centre, which serves approximately 15,000 people. Mapanza is 75 kilometers from the Southern Province provincial capital of Choma and is a rural area comprised of villages of multi- and single-structure homesteads often housing an extended family. Rearing and keeping livestock is common in the region. The rainy season is from November to April, while the cool-dry season is from May to August and the hot-dry season from August to October. In 2018, when this study was conducted, malaria prevalence by rapid diagnostic test in children younger than five years of age was 0.9%, and most infections occurred in April just after the rainy season (18).

The study was an open-enrollment prospective longitudinal cohort of geographically contiguous households. Each study month, 202 households were surveyed for an epidemiological study and a subset of 59 (29%) households was randomly selected and visited each month for entomological surveillance. Entomological households were purposefully selected based on their proximity to expected larval sources At each visit, information was collected on the household construction, animal ownership, modes of transportation, household amenities, and the receipt of malaria interventions such as IRS, mass drug administration (MDA), or reactive test and treat. In all households selected for entomological surveillance, two CDC miniature light traps (John W. Hock Co., Gainesville) were set, one indoors and outdoors, 1.5 meters above the ground in the sleeping area and near outdoor gathering areas on the porch of the house. If households had domestic animals nearby, a third trap was set near the animal pen.

Traps were run from 18:00 to 6:00 the following morning. Field teams recorded the location of the mosquito trap (indoor versus outdoor and proximity to people or animals), and physical conditions of the structure, and the number of bednets hanging and IRS history. Mosquitoes were killed by freezing and morphologically identified and enumerated as either anopheline or culicine. If anopheline, morphological identification was completed to species and specimens individually stored on silica. These processes have been previously described (19,20). All anopheline mosquitoes were split into head/thorax and abdomen. Abdomens were homogenized and DNA was extracted using a modified salt extraction protocol (21). Anopheline identities were confirmed by PCR using diagnostic tools designed for the *An. gambiae* complex, the *An. funestus* complex, and a third targeting the internal transcribed spacer 2 (ITS2) region of nuclear rDNA (22–25). Enzyme-linked immunosorbent assay (ELISA) analysis was used to detect the circumsporozoite protein (CSP) of *Plasmodium falciparum* in the head/thorax all anophelines, and samples ELISA-positive for CSP were confirmed to contain *P. falciparum* DNA using qPCR (8).

### Environmental and spatial data

Household location data was collected using GPS enabled tablets during each household visit, and coordinate locations were mapped in ArcGIS Pro version 2.8.3 (ESRI, Redlands, Canada). Environmental features were extracted in ArcGIS through supervised classification of an adjusted orthomosaic color balance from 2-meter resolution multispectral satellite imagery. Leveraging contrasts in spectral bands, a classification scheme was created to distinguish land types (Figure 1). The ISO Cluster Unsupervised Classification tool in ArcGIS was used to classify land type before samples were defined to train the classification tool to be more accurate. The percentage of land type was calculated within 50 m buffers around each household to represent likely resting habitat of mosquitoes feeding at the household. Distance from households to the nearest bodies of water in meters was calculated. Bodies of water include streams, rivers, and any land type classified as water that is greater than 20 square meters in area. Temperature and precipitation data were collected using a HOBO Micro Station (Onset Computer Corporation, Bourne, MA) set in the study area.

**Figure 1.**
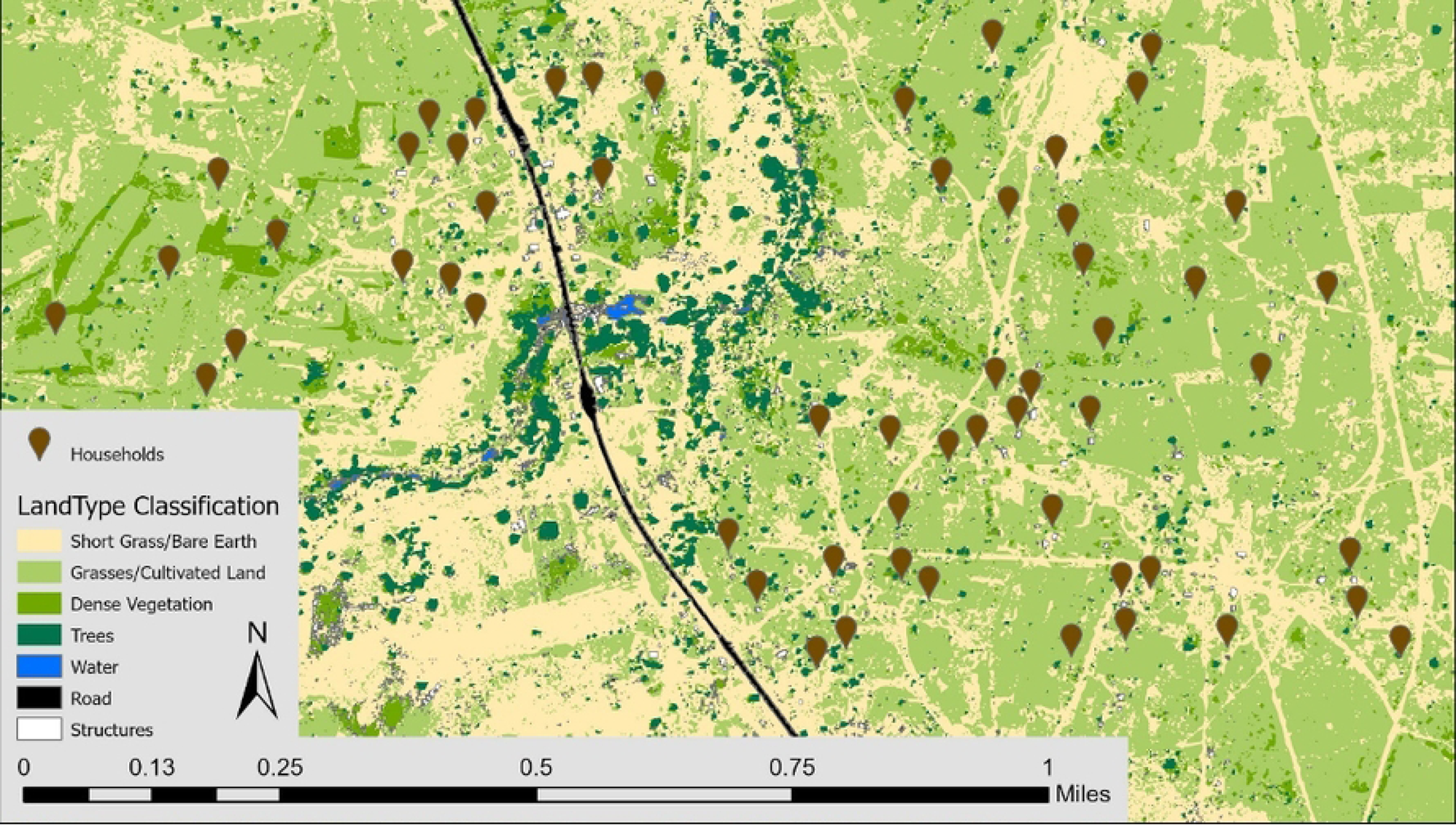
Land type cover map of Mapanza, Choma District, Zambia featuring participating households.

### Statistical analysis

Generalized linear mixed effects negative binomial regression with zero-inflation models were used to describe the relationship between risk factors and the outcome of monthly anopheline counts in each trap at each household. Six separate models were run after stratifying the data by season (all, rainy, and dry) and trap location (indoor and outdoor on porch), to allow for the detection of different risk factors at different levels of importance. There were too few observations to run regression analyses on outdoor traps set near animal pens. Household random effects were added to each model to test if within-household correlation over time contributed to explaining variability. The random effect was included in each of the final models if a likelihood ratio test comparing each model with and without the random effect was significant at an alpha of 0.10. The zero-inflation model accounted for the high number of zero-count traps and included covariates of temperature, rain, number of people sleeping inside, and time spent outside. Risk factors considered for inclusion were drawn from an a priori constructed conceptual framework highlighting the theorized causal relationship of mosquito count by chemosensory cues such as animal or human factors, chemical (insecticides) and non-chemical (burning fire) deterrents, seasonality, climate, and environmental suitability. Variables were included in the final multivariable regression models if they were significant in univariable testing at an alpha level of 0.10; households were included in the final multivariable regression if they contained data for all variables selected as risk factors. Analysis was conducted in R version 4.1.1 and ArcGIS Pro version 2.8.3. Two sensitivity analyses were performed. First, the risk factor analysis was repeated using the outcome of all mosquitoes (anophelines and culicines) to offer a comparison between anopheline and all mosquitoes. Second, the risk factor analysis was repeated using the outcome of *An. arabiensis* only, to isolate risk factors associated with a known malaria vector.

## Results

A total of 2,364 traps were set for 1,130 household collection events from 59 households between October 2018 and September 2020, although data collection was paused from April to June of 2020 due to the COVID-19 pandemic. Of these, 1,121 traps were set indoors and 1,243 traps were set outdoors (1,032 on household porch and 211 near animal pens) (Table 1). Approximately 30% of households owned at least one goat. Brick and iron sheets were the most common respective wall and roof materials, 52% of houses had open eaves, and 79% of households had holes in structure walls. At the time of each visit, households had less than one bednet hanging on average and 18% reported IRS in the previous 12 months. Active use of bednets and recent IRS were more common in rainy season than dry season. Fires were burned near 81% of outdoor and 14% of indoor traps and were more common in the dry season. As expected, rain and temperature differed by season, with average daily temperatures of 23.7 and 27.7 C and total biweekly rainfall of 1.3 and 40.0 millimeters of rain in the dry and rainy seasons, respectively. Land type characteristics did not differ by season.

**Table 1.**
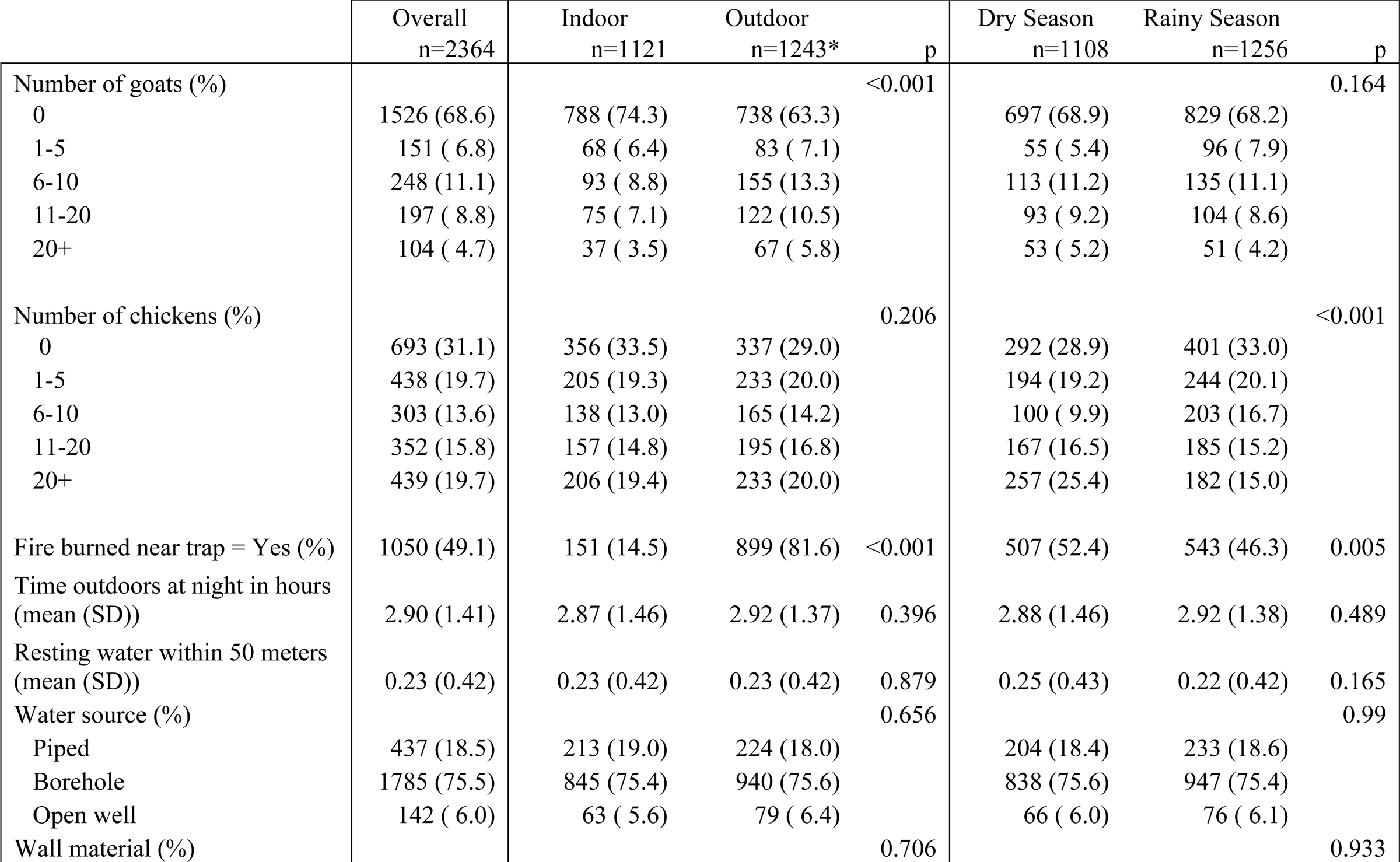
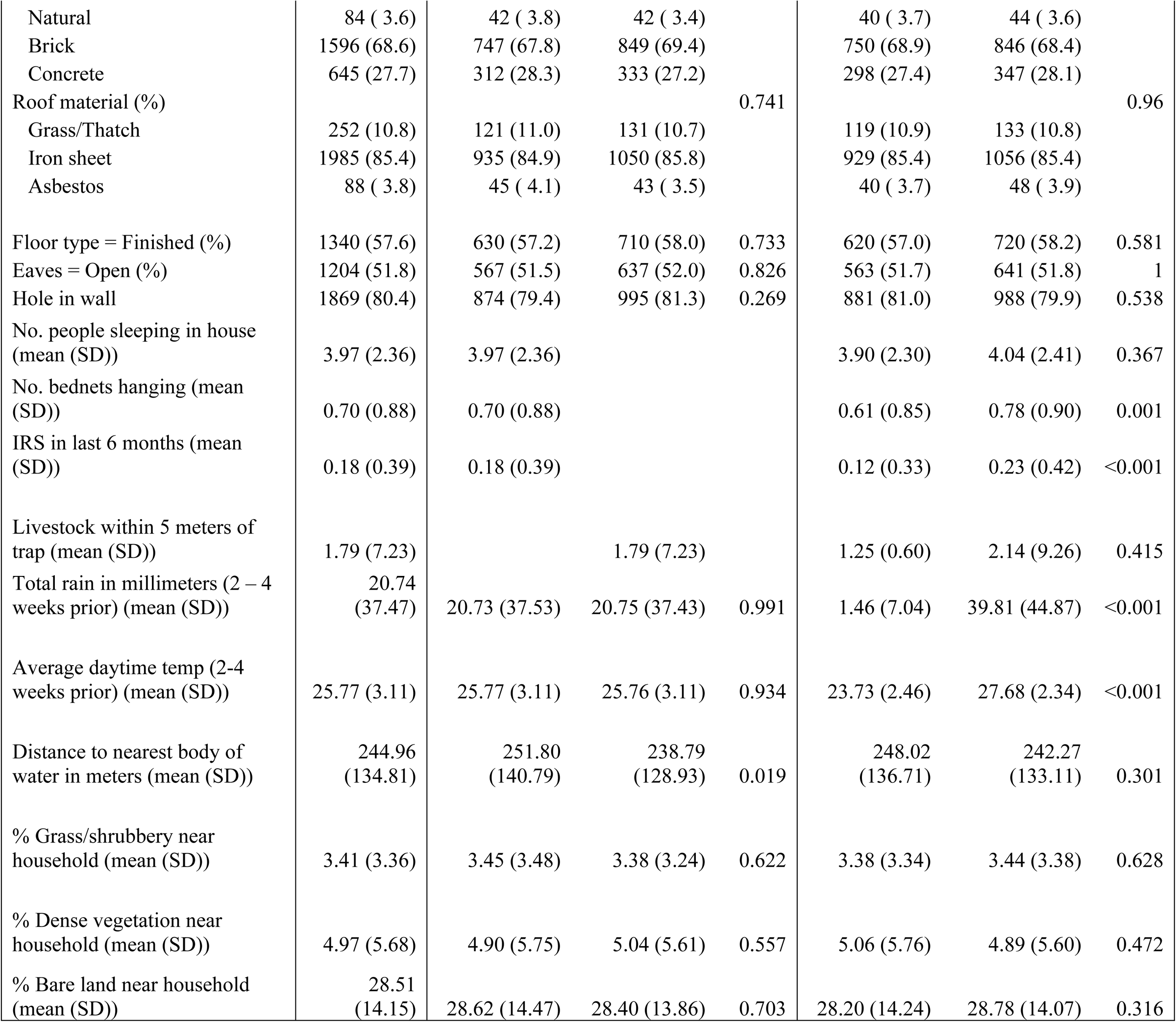
Baseline characteristics of household collections by season and trap location. P-values were calculated via the Wald and chi-squared methods for continuous and categorical variables, respectively.

|  | Overall<br>n=2364 | Indoor<br>n=1121 | Outdoor<br>n=1243* | p | Dry Season<br>n=1108 | Rainy Season<br>n=1256 | p |
| --- | --- | --- | --- | --- | --- | --- | --- |
| Number of goats (%) |  |  |  | <0.001 |  |  | 0.164 |
| 0 | 1526 (68.6) | 788 (74.3) | 738 (63.3) |  | 697 (68.9) | 829 (68.2) |  |
| 1-5 | 151 ( 6.8) | 68 ( 6.4) | 83 ( 7.1) |  | 55 ( 5.4) | 96 ( 7.9) |  |
| 6-10 | 248 (11.1) | 93 ( 8.8) | 155 (13.3) |  | 113 (11.2) | 135 (11.1) |  |
| 11-20 | 197 ( 8.8) | 75 ( 7.1) | 122 (10.5) |  | 93 ( 9.2) | 104 ( 8.6) |  |
| 20+ | 104 ( 4.7) | 37 ( 3.5) | 67 ( 5.8) |  | 53 ( 5.2) | 51 ( 4.2) |  |
| Number of chickens (%) |  |  |  | 0.206 |  |  | <0.001 |
| 0 | 693 (31.1) | 356 (33.5) | 337 (29.0) |  | 292 (28.9) | 401 (33.0) |  |
| 1-5 | 438 (19.7) | 205 (19.3) | 233 (20.0) |  | 194 (19.2) | 244 (20.1) |  |
| 6-10 | 303 (13.6) | 138 (13.0) | 165 (14.2) |  | 100 ( 9.9) | 203 (16.7) |  |
| 11-20 | 352 (15.8) | 157 (14.8) | 195 (16.8) |  | 167 (16.5) | 185 (15.2) |  |
| 20+ | 439 (19.7) | 206 (19.4) | 233 (20.0) |  | 257 (25.4) | 182 (15.0) |  |
| Fire burned near trap = Yes (%) | 1050 (49.1) | 151 (14.5) | 899 (81.6) | <0.001 | 507 (52.4) | 543 (46.3) | 0.005 |
| Time outdoors at night in hours<br>(mean (SD)) | 2.90 (1.41) | 2.87 (1.46) | 2.92 (1.37) | 0.396 | 2.88 (1.46) | 2.92 (1.38) | 0.489 |
| Resting water within 50 meters<br>(mean (SD)) | 0.23 (0.42) | 0.23 (0.42) | 0.23 (0.42) | 0.879 | 0.25 (0.43) | 0.22 (0.42) | 0.165 |
| Water source (%) |  |  |  | 0.656 |  |  | 0.99 |
| Piped | 437 (18.5) | 213 (19.0) | 224 (18.0) |  | 204 (18.4) | 233 (18.6) |  |
| Borehole | 1785 (75.5) | 845 (75.4) | 940 (75.6) |  | 838 (75.6) | 947 (75.4) |  |
| Open well | 142 ( 6.0) | 63 ( 5.6) | 79 ( 6.4) |  | 66 ( 6.0) | 76 ( 6.1) |  |
| Wall material (%) |  |  |  | 0.706 |  |  | 0.933 |
| Natural | 84 ( 3.6) | 42 ( 3.8) | 42 ( 3.4) |  | 40 ( 3.7) | 44 ( 3.6) |  |
| Brick | 1596 (68.6) | 747 (67.8) | 849 (69.4) |  | 750 (68.9) | 846 (68.4) |  |
| Concrete | 645 (27.7) | 312 (28.3) | 333 (27.2) |  | 298 (27.4) | 347 (28.1) |  |
| Roof material (%) |  |  |  | 0.741 |  |  | 0.96 |
| Grass/Thatch | 252 (10.8) | 121 (11.0) | 131 (10.7) |  | 119 (10.9) | 133 (10.8) |  |
| Iron sheet | 1985 (85.4) | 935 (84.9) | 1050 (85.8) |  | 929 (85.4) | 1056 (85.4) |  |
| Asbestos | 88 ( 3.8) | 45 ( 4.1) | 43 ( 3.5) |  | 40 ( 3.7) | 48 ( 3.9) |  |
| Floor type = Finished (%) | 1340 (57.6) | 630 (57.2) | 710 (58.0) | 0.733 | 620 (57.0) | 720 (58.2) | 0.581 |
| Eaves = Open (%) | 1204 (51.8) | 567 (51.5) | 637 (52.0) | 0.826 | 563 (51.7) | 641 (51.8) | 1 |
| Hole in wall | 1869 (80.4) | 874 (79.4) | 995 (81.3) | 0.269 | 881 (81.0) | 988 (79.9) | 0.538 |
| No. people sleeping in house (mean (SD)) | 3.97 (2.36) | 3.97 (2.36) |  |  | 3.90 (2.30) | 4.04 (2.41) | 0.367 |
| No. bednets hanging (mean (SD)) | 0.70 (0.88) | 0.70 (0.88) |  |  | 0.61 (0.85) | 0.78 (0.90) | 0.001 |
| IRS in last 6 months (mean (SD)) | 0.18 (0.39) | 0.18 (0.39) |  |  | 0.12 (0.33) | 0.23 (0.42) | <0.001 |
| Livestock within 5 meters of trap (mean (SD)) | 1.79 (7.23) |  | 1.79 (7.23) |  | 1.25 (0.60) | 2.14 (9.26) | 0.415 |
| Total rain in millimeters (2 – 4 weeks prior) (mean (SD)) | 20.74 (37.47) | 20.73 (37.53) | 20.75 (37.43) | 0.991 | 1.46 (7.04) | 39.81 (44.87) | <0.001 |
| Average daytime temp (2-4 weeks prior) (mean (SD)) | 25.77 (3.11) | 25.77 (3.11) | 25.76 (3.11) | 0.934 | 23.73 (2.46) | 27.68 (2.34) | <0.001 |
| Distance to nearest body of water in meters (mean (SD)) | 244.96 (134.81) | 251.80 (140.79) | 238.79 (128.93) | 0.019 | 248.02 (136.71) | 242.27 (133.11) | 0.301 |
| % Grass/shrubbery near household (mean (SD)) | 3.41 (3.36) | 3.45 (3.48) | 3.38 (3.24) | 0.622 | 3.38 (3.34) | 3.44 (3.38) | 0.628 |
| % Dense vegetation near household (mean (SD)) | 4.97 (5.68) | 4.90 (5.75) | 5.04 (5.61) | 0.557 | 5.06 (5.76) | 4.89 (5.60) | 0.472 |
| % Bare land near household (mean (SD)) | 28.51 (14.15) | 28.62 (14.47) | 28.40 (13.86) | 0.703 | 28.20 (14.24) | 28.78 (14.07) | 0.316 |

Of the 2,364 traps set, 1,525 (65%) caught zero mosquitoes, and 1,960 (83%) caught zero female anophelines. In total, 3,218 mosquitoes were collected, of which 1,532 were female anophelines. Across all seasons, fifty-five percent of female anophelines were collected outdoors by animal traps, 21% were collected outdoors by people on porches, and 24% were collected indoors. This distribution shifted during the dry season when there were fewer anophelines overall and only 11% were caught indoors whereas 24% were caught outdoors on porches near where people gather (Figure 2). Through morphological and subsequent molecular analysis, ten species of anophelines were identified, the largest proportion of which were *An. arabiensis*, comprising 80% of anophelines collected indoors and 28% of those collected outdoors across porch and animal pen traps (Figure 3). The second most abundant anopheline was *An. quadriannulatus,* followed by *An. rufipes* (Figure 3) which were most commonly found by outdoor animal pens. Notably, *An. rufipes* was also the most abundant vector found near human gathering spaces outdoors during dry season. Of all anophelines, 6% (78) produced a 500 bp band with the ITS2 PCR assay, identifying them as either *An. rufipes*, *An. maculipalpis* or *An. pretoriensis*.(22) *Anopheles arabiensis* and *An. squamosus* had higher abundances in the rainy season, while *An. leesoni, An. pretoriensis, An. quadriannulatus*, and *An. rufipes* were more abundant in the dry season (p-values all <0.05, Table S11). Total *An. arabiensis* abundance did not differ in indoor collections versus outdoor collections, (p-value =0.362, Table S2) although all other species (p-values all <0.001, Table S1b) were collected more frequently outdoors. There was one *P. falciparum* parasite-positive mosquito, an *An. squamosus* collected outdoors.

**Figure 2.**
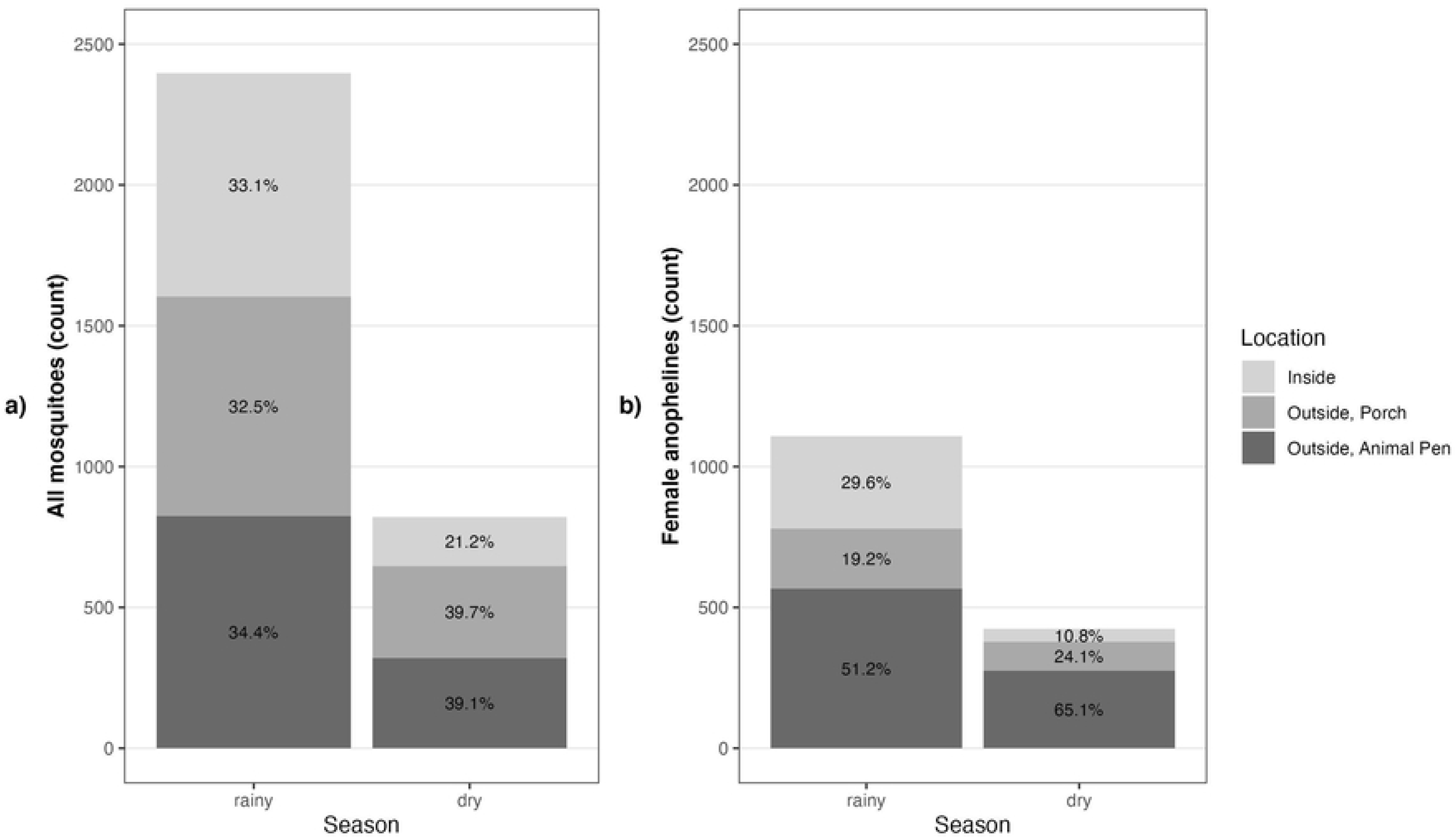
Mosquito counts from all traps by season and collection location. Panel a shows the number of all mosquitoes (anophelines and culicines) and panel b shows the number of female anophelines.

**Figure 3.**
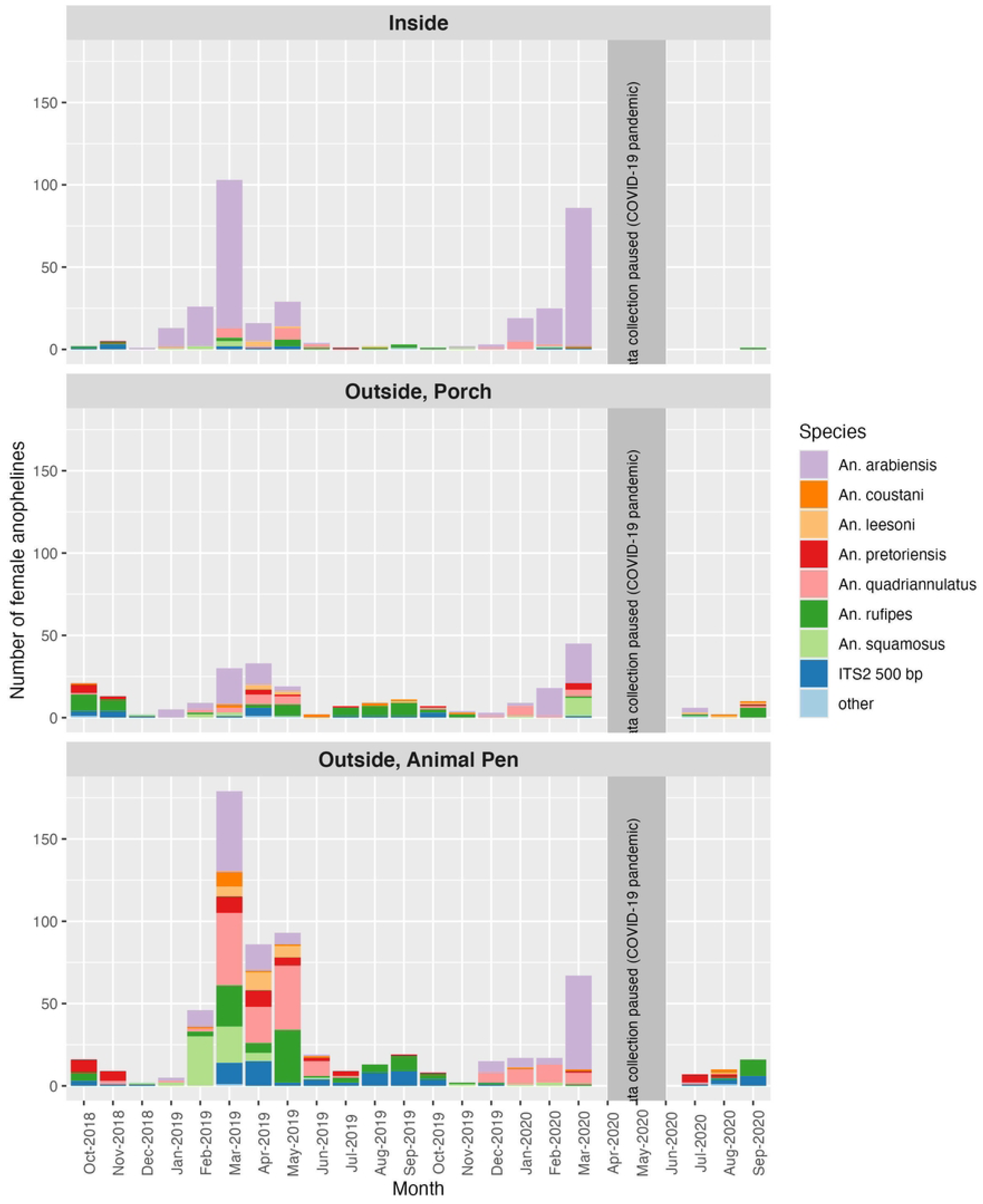
Number of female anophelines by species per month by collection location. Samples identified in the legends as ITS2 500 bp may be either *An. rufipes*, *An. maculipalpis* or *An. pretoriensis*. There were a total of seven sample identified as either *Anopheles longipalpis* (n=4), *An. rivulorum* (n=1) and *An. rivulorum-like* (n=2), and these are grouped together as ‘other’.

Results of the multivariable regressions are plotted in Figure 4, and tables S2 – S7 contain the full results of the univariable models and each of the six multivariable regression models. The household random effect significantly contributed to explaining model variance only in the indoor all season and indoor rainy season models. Since separate regressions were run for the dry season, rainy season, and for both seasons combined, the risk factors significant in univariable analysis and included in the multivariable analysis and significant in the multivariable analysis, differed. There were no common risk factors associated with indoor anopheline abundance across dry and rainy seasons. In the dry season, owning more goats (11 – 20 goats), and each additional centimeter of rain were associated with 3.5 (IRR 4.51, 95% confidence interval [CI]: 3.49, 5.84), and 1.4 (IRR 2.35, 95% CI: 2.21, 2.50) times more indoor anophelines. In the rainy season, each additional bednet hanging and having holes in the wall were associated with 0.64 (IRR 1.64, 95% confidence interval [CI]: 1.16 2.32) and 1.6 (IRR 2.57, 95% CI: 1.08, 6.07) times higher indoor anopheline counts. While in the dry season, each single degree increase in temperature was associated with 1.11 (IRR 2.11, 95% CI: 1.44, 3.10) times more indoor anopheline mosquitoes, during the rainy season each single degree increase in temperate was associated with 0.27 (IRR 0.73, 95% CI: 0.59, 0.90) times fewer indoor anopheline. Eave type (open or closed) was not associated with change in anopheline counts in either season, although holes in the wall were associated with a 1.57 (IRR 2.57, 95% CI: 1.08, 2.07) times increase in indoor anophelines in rainy season only. Having a finished floor was also associated with a 0.50 (IRR 0.50, 95% CI: 0.28, 0.87) times decrease in indoor anophelines, though in rainy season only.

**Figure 4.**
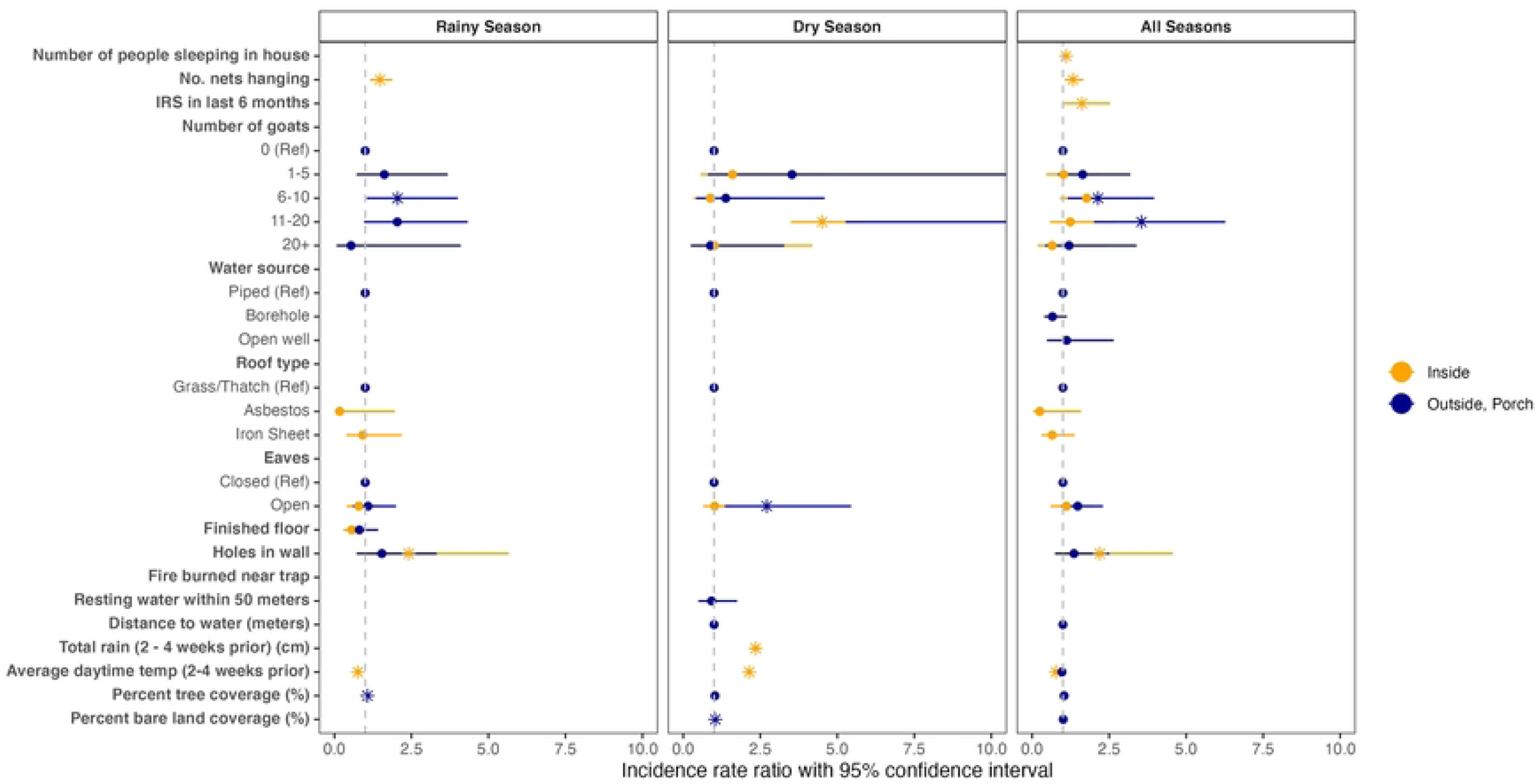
Risk factors associated with indoor and outdoor anopheline counts in the dry and rainy seasons. Estimates were generated through multivariable regression. Asterisks indicate those variables that are statistically significant at an alpha of 0.05.

There were several risk factors associated with anophelines caught outdoors in traps set on the porch where humans gather. In the dry season, increasing goat ownership (11 - 20 goats), open eaves, and each percent increase of bare land cover were associated with 12 (IRR 13.19, 95% CI:5.27, 33.03), 1.7 (IRR 2.71, 95% CI: 1.35, 5.45) and 0.04 (IRR 1.04, 95% CI: 1.01 - 1.07) times higher incidence of anophelines. In the rainy season, increasing goat ownership (6 - 10 goats) and each percent increase of tree cover increase were associated with 1.0 (IRR 2.05, 95% CI: 1.05, 4.01) and 0.07 (IRR 1.07, 95% CI: 1.01 - 1.12) times higher incidence of anophelines. The *An. arabiensis* sensitivity analysis only found goat ownership to be a significant risk factor for outdoor rainy season *An. arabiensis*, confirming that this association is not exclusively due to an increase of zoophilic non-malaria vectors (Table S16). The sensitivity analysis assessing risk factors for all mosquitoes uncovered similar associations in indoor and outdoor mosquitoes, although there were no associations between IRS or bednets hanging (Figure S1, Tables S8 – S13). Full results of the *An. arabiensis* and all mosquito sensitivity analyses are in the supplement (Tables S14 – S19).

## Discussion

Relatively few studies have examined risk factors of anopheline abundance in low malaria transmission areas, even though residual transmission is dependent on the presence of local vectors. Here, we found evidence to support our hypothesis that hyperlocal land type is associated with outdoor vector abundance and we identified strong seasonal patterns in indoor and outdoor vector species and abundance that have implications for transmission and vector control strategies. First, all vectors were much more abundant during the rainy season, during which lower quality housing and animal ownership were risk factors for indoor and outdoor vectors respectively. Second, *An. arabiensis* was the most common indoor and outdoor anopheline species. Even in the absence of any CSP-positive *An. arabiensis*, its high indoor abundance during the rainy season, relatively high abundance around human gathering places outdoors during rainy season, highly anthropophilic feeding, and high competence as a vector suggest it is the primary vector driving rainy-season transmission (26). Thus, its control remains important in this area. Third, outdoor anophelines were more abundant than indoor vectors, and proportionately much more abundant in the dry season when *An. arabiensis* and indoor vectors all but disappeared. These outdoor dry season anophelines are understudied species, largely considered zoophilic but with significant behavioral plasticity as opportunistic human feeders (7–9). Other pre-elimination settings, in Ethiopia and Kenya have similarly found a predominance of outdoor vectors, including sporozoite positive *An. coustani*, *An. longipalpis*, and *An. rivulorum* (27,28). Here, the only *P. falciparum*-positive mosquito was outdoor-caught *An. squamosus*, which reinforces the long-term albeit infrequent reports of *P. falciparum* CSP-positive *An. squamosus* from in this area (29). *An. rufipes* has been noted as an important secondary vector elsewhere in Zambia, and is the dominant dry-season species caught in this study (30). Recent genotyping of *P. falciparum* infections across multiple rainy seasons suggested that transmission persists through dry-season; *An. rufipes* and similar dry-season vectors are thus implicated in this transmission and therefore their control is essential to to achieviing elimination (17). Control strategies, therefore, need to target vectors year-round, expand to target exophagic and zoophilic mosquitoes, and leverage known risk factors when selecting methods of control.

During the rainy season, indoor vector control remains important. Increases in indoor anopheline counts were associated with lower housing quality and having more bednets hanging. There was not an association between all indoor mosquito (anopheline and culicine) counts and utilizing bednets, suggesting this behavior change could also be related to risk perception in addition to perception of increased biting of all mosquitoes, not just anophelines. Housing modifications, including covering holes in walls, may also reduce indoor anopheline counts in the rainy season and have been shown to be associated with decreased malaria risk (13,31,32). Outdoor vector control is important to consider in both rainy and dry seasons, the former to curb outbreaks and the latter to possibly interrupt residual transmission. Individuals are unprotected from bites received outdoors, where we caught the majority of anophelines in both rainy and dry seasons. Outdoor vectors were associated with increased goat ownership and using an open well in all seasons. There are currently few options for outdoor vector control, although novel methods are under development. Recent trials of attractive toxic sugar baits (ATSB) in Southern and Western Zambia did not show reductions in mosquito parity or abundance (33). There is promise in spatial repellant devices, which vary by design, but may be hung in outdoor areas where people congregate, and when impregnated with insecticide, reduce vector abundance (34,35). We should not dismiss household level activities and behaviors that can reduce outdoor abundance in the dry season, e.g., fires burning near people, covering open wells, and managing additional environmental anopheline breeding locations which may appear at the outset of the rainy season. Applying larvicide to outdoor breeding sites has had some success in reducing biting and vector populations in some settings but the logistics of large scale and frequent applications are challenging (36). However, this might be feasible in the dry season if productive sites can be identified and are nearby households (35,36). The increased risk of vector abundance for families who keep animals should be considered as well, and solutions including zooprophylaxis and the use livestock as a mode of vector control, i.e. applying insecticide onto cattle via sponge or subcutaneous implant, have long been discussed (37).

This work is limited in that it cannot directly connect these entomological patterns with local malaria epidemiology. We do not report on malaria incidence and only detected one sporozoite-positive mosquito. We must then infer the impact of these trends in anopheline abundance on malaria risk. Additionally, we assume that CDC light trap collections capture mosquitoes seeking a human blood meal, though this may not always be the case. However, our sensitivity analysis that examined abundance of *An. arabiensis*, which is highly anthropophilic, assures us that these risk factors are associated with vectors that will seek human hosts. Alternative collection methods such as human landing catches may have produced more accurate estimates of human biting but would have reduced the overall number of collections. Human landing catches are planned in future studies in this area. Further, the number of traps and anophelines caught provide adequate power to identify risk factors across collection locations and seasons, which is a major strength of this analysis, as the season-specific species and risk factors findings would otherwise be obscured if the analysis was not so stratified.

This study provides a thorough and important profile of anopheline species distribution and abundance in an area targeting malaria elimination but hampered by continued residual transmission. The seasonal stratification of vector species and the finding that dry season anophelines are largely understudied species that exhibit outdoor behaviors, clarifies the need to deliver vector control during the dry season to interrupt cross-seasonal transmission. Typically, concerted vector control efforts for malaria are concentrated during the rainy season but elimination or reduction of these dry season anophelines may be critical to interrupting transmission in this ecological setting. This study also exemplifies the importance of continued vector surveillance, which may be reactive in areas with the occasional case in dry season, in areas approaching elimination to inform vector control in areas of seasonal and low but persistent transmission.

## Ethics approval and consent to participate

The study was approved by the Zambian Tropical Diseases Research Center (TDRC under IRB no: TDRC/ERC/2010/14/11 and the Johns Hopkins Bloomberg School of Public Health (Baltimore, Maryland) under IRB no: 00003467.

## Consent for publication

Not applicable.

## Availability of data and materials

Under the National Health Research Act, the Government of Zambia does not allow public access to data collected in Zambia. All investigators interested in the datasets supporting the conclusions of this article are required to submit a written request to the Ministry of Health. Contact the Macha Research Trust IRB Chairperson (, +260979402560) to coordinate the request.

## Author contributions

ACM conducted analysis and wrote the final manuscript. VK conceived of and carried out the initial analytic plan and produced an early draft. LS, TM, HH lead data specimen collection and laboratory processing. MEG and RLMNA conducted additional laboratory processing and verification. TS, MRD, and FCC supported design and and contributions to spatial analysis. JLS, JCS, WJM, and DEN conceived of the parent study and managed and oversaw data collection. All authors contributed to and reviewed the final manuscript.

## Acknowledgements

The authors gratefully acknowledge all participants and the Zambian communities of Choma District. The authors thank the National Malaria Elimination Centre for their support and all ANTOOMWE study team members and study participants.

## Supporting information

**Figure S1**. **Risk factors associated with indoor and outdoor total mosquitoes in dry and rainy season.** Estimates were generated through multivariable regression; asterisks indicate those variables that are statistically significant at an alpha of 0.05. Corresponding full regression outputs can be found in Tables S8 – S13.

**Table S1a**. **Count of anophelines by species and season for October 2018 – September 2019.** The counts were restricted to these dates given the study was paused for three months of the 2020 dry season (April – June 2020) due to the Covid-19 pandemic. P-values were calculated using the chi-squared test.

**Table S1b. Count of anophelines by collection location for the entire study period.** P-values were calculated using the chi-squared test.

**Table S2. Multivariable regression for risk factors associated with indoor anopheline abundance in the dry season.**

**Table S3. Multivariable regression for risk factors associated with outdoor (porch collection) anopheline abundance in the dry season.**

**Table S4. Multivariable regression for risk factors associated with indoor anopheline abundance in the rainy season.**

**Table S5. Multivariable regression for risk factors associated with outdoor (porch collection) anopheline abundance in the rainy season.**

**Table S6. Multivariable regression for risk factors associated with indoor anopheline abundance across all seasons.**

**Table S7. Multivariable regression for risk factors associated with outdoor (porch collection) anopheline abundance across all seasons.**

**Table S8. Multivariable regression for risk factors associated with indoor mosquito abundance in the dry season.**

**Table S9. Multivariable regression for risk factors associated with outdoor (porch collection) mosquito abundance in the dry season.**

**Table S10. Multivariable regression for risk factors associated with indoor mosquito abundance in the rainy season.**

**Table S11. Multivariable regression for risk factors associated with outdoor (porch collection) mosquito abundance in the rainy season.**

**Table S12. Multivariable regression for risk factors associated with indoor mosquito abundance across all seasons.**

**Table S13. Multivariable regression for risk factors associated with outdoor (porch collection) mosquito abundance across all seasons.**

**Table S14. Multivariable regression for risk factors associated with indoor *An. arabiensis* abundance in the dry season.**

**Table S15. Multivariable regression for risk factors associated with outdoor (porch collection) *An. arabiensis* abundance in the dry season.**

**Table S16. Multivariable regression for risk factors associated with indoor *An. arabiensis* abundance in the rainy season.**

**Table S17. Multivariable regression for risk factors associated with outdoor (porch collections) *An. arabiensis* abundance in the rainy season.**

**Table S18. Multivariable regression for risk factors associated with indoor *An. arabiensis* abundance across all seasons.**

**Table S19. Multivariable regression for risk factors associated with outdoor (porch collection) *An. arabiensis* abundance across all seasons.**

